# An initial validation study of DigiBel, a web-application enabling self-assessment of air and bone-conduction audiometry in the community

**DOI:** 10.1101/2023.04.11.23288179

**Authors:** Anna Sienko, Arun J Thirunavukarasu, Tanya Kuzmich, Louise E Allen

## Abstract

80% of primary school children suffer from glue ear which may impair hearing at a critical time for speech acquisition and social development. An online application, DigiBel, has been developed primarily to identify individuals with conductive hearing impairment who may benefit from temporary use of bone-conduction (BC) assistive technology in the community. This preliminary study aims to determine the screening accuracy and usability of DigiBel self-assessed air-conduction (AC) pure tone audiometry (PTA) in adult volunteers with simulated hearing impairment prior to formal clinical validation.

Healthy adults, each with one ear plugged, underwent standard automated AC PTA (reference test) and DigiBel audiometry in quiet community settings. Threshold measurements were compared across six tone frequencies and DigiBel test-retest reliability was calculated. The accuracy of DigiBel for detecting more than 20 decibels of hearing impairment was assessed.

30 adults (30 unplugged ears and 30 plugged ears) completed both audiometry tests. DigiBel had 100% sensitivity (95%CI 87.23-100) and 72.73% (95%CI 54.48-86.70) specificity in detecting hearing impairment. Threshold mean bias was insignificant except at 4000 and 8000Hz where a small but significant over-estimation of threshold measurement was identified. All 24 subjects completing feedback rated the DigiBel test good or excellent and 21(87.5%) agreed or strongly agreed that they would be able to do the test at home without help.

This study supports the potential use of DigiBel as a screening tool for hearing impairment. The findings will be used to improve the software further and undertake a formal clinical trial of AC and BC audiometry in individuals with suspected conductive hearing impairment.

**Author Summary:** Hearing loss is a major global health issue. It can affect many aspects of life such as education, employment, communication, and result in social isolation. Two thirds of people with severe hearing loss live in low and middle countries with poor access both to hearing testing (audiometry) or conventional hearing aids.

Several software applications (apps) like DigiBel, studied here, have been developed to enable individuals to test their own hearing in the community. Uniquely, DigiBel has the additional potential to identify individuals with hearing loss who could derive immediate hearing support from an affordable and rechargeable bone-conduction headphone / microphone kit while waiting specialist care.

This initial study of DigiBel provides confirmation that the app is easy to use and accurate at detecting simulated hearing impairment. It lays the groundwork for future clinical studies to assess DigiBel’s performance in children and adults with hearing impairment.

## Introduction

Over 5% of the world’s population, approximately 430 million people worldwide, have disabling hearing loss; 34 million of these are children. (1, 2) Children in low and middle income countries, families experiencing socio-economic deprivation and disadvantaged populations are disproportionally affected by conductive hearing loss caused by glue ear (otitis media with effusion) and its complications, or from damage to the ear drum as seen in chronic tympanic perforations or chronic serous otitis media. (3, 4) Since delayed recognition and management of childhood hearing impairment has long-term consequences for socialisation and educational attainment, screening audiometry is recommended in primary school. (5) However, population screening programmes are hindered by cost, standardisation, requirement for staff-training, false-positive referrals, and poor data capture. (6, 7) Even where available, school screening may miss children with fluctuating hearing loss due to glue ear. Additionally, several year groups have missed screening during the Covid pandemic. These children may face months of impaired hearing before diagnosis and management due to backlogs in audiometry and specialist services.

Hearing thresholds are assessed using pure-tone audiometry (PTA) with air-conduction (AC) and bone-conduction (BC) headphones, but this usually requires specialist equipment and trained clinicians. Automated audiometry and, more recently, validated self-testing hearing software applications (apps), may improve the accessibility of screening and threshold audiometry testing, particularly in rural areas. DigiBel is a recently developed CE marked online app which enables self-testing of AC and BC hearing levels. Like some other audiometry apps, it is suitable for community use in adults and children without clinical support. Its potential, more unique, function is to identify children with hearing impairment who may derive functional benefit from using a BC headphone / microphone kit (Raspberry Pi, Cambridge, UK) while waiting for diagnosis, spontaneous resolution or definitive management of their glue ear (8).

The purpose of this preliminary study of DigiBel audiometry, undertaken in healthy adults situated in quiet community settings, is to determine the app’s sensitivity and specificity for detecting simulated conductive hearing impairment of more than 20 decibels (dB). The secondary aim is to assess the web-application’s usability to enable improvement of the app design.

## Materials and methods

In this comparative study, healthy adult volunteers from the community without previous history of hearing impairment were invited to participate. After receiving an explanation of the study, participants provided verbal consent to proceed to audiometry testing. Each participant was assigned a unique study identification number; no personal identifiable information was recorded. The study adhered to the tenets of the Declaration of Helsinki and the protocol was approved by the Quality and Safety Committee of Cambridge University Hospitals NHS Foundation Trust as part of a service improvement project.

Testing was undertaken in community settings such as participants’ homes and classrooms. Prior to testing, each volunteer placed a foam earplug into their left ear to simulate a conductive hearing loss, making each ear an independent entity for the purpose of statistical analysis and providing a range of hearing levels.

The reference automated AC PTA and the index DigiBel audiometry test were undertaken one after the other in random sequence. DigiBel audiometry for four frequencies was repeated immediately after the initial test, to assess test-retest (TRT) variability. After completing both audiometry tests, each volunteer was asked to complete a feedback questionnaire covering test preference and usability.

### Automated Pure Tone audiometry—the gold-standard reference test

Automated PTA was undertaken using an Oscilla (Oscilla A/S Aarhus Denmark) USB300 audiometer with TDH-39 headphones. The modified Hughson-Westlake algorithm was used for determining the reference AC audiometric threshold. (9). After an initial explanation by the technician and a conditioning test at 1000Hz, thresholds were recorded at 2000, 4000, 8000, 1000, 500, then 250Hz in accordance with British Society of Audiology recommendations (10). The hearing threshold criterion for each frequency was determined as the lowest intensity at which participants accurately signalled two confirmations out of three presentations. The number of false positive responses was manually recorded.

### The DigiBel index test

DigiBel has been laboratory and biologically calibrated (Institute of Sound and Vibration Research, University of Southampton, UK and Chears-audiology, Royston, UK) to run specifically on any model iPad tablet (Apple, Cupertino, California, USA) with Seinnheiser HD 400S AC headphones (Wedemark, Germany). BC audiometry (not tested in this study) uses Bluetooth-connected Raspberry Pi children’s BC headphones.

A pre-test embedded video provides instructions for use and a checklist ensures that the iPad volume is on maximum and the headphones are fitted correctly. Noise sampling, using the inbuilt Sennheiser headphone microphone, ensures that the ambient noise level is less than 40 decibels (dB) prior to testing. Configurable settings include a choice of warble / pure tone, test frequencies, ambient noise setting and child or adult version of the test.

The user taps a central animated button on the iPad display when they hear the tone (Fig 1A). A conditioning step requires the user to accurately tap the button on hearing a random onset supra-threshold tone at 1000Hz before testing can begin. During testing, the onset of the tone is randomised from 0-3 seconds after appearance of the response button to avoid a predictable response pattern. The tone stops in response to the tap and a psychophysical staircase algorithm starting from 60dB (10dB down, 5dB up) is followed requiring three reversals dependent on the user’s input. The final threshold is calculated by the mean of the three reversal thresholds. Once completed, a standard audiometry graph and the number of false positive responses is displayed (Fig 1B). The user can choose to repeat testing or undertake BC testing to determine the functional effect on hearing levels.

**Fig 1.**
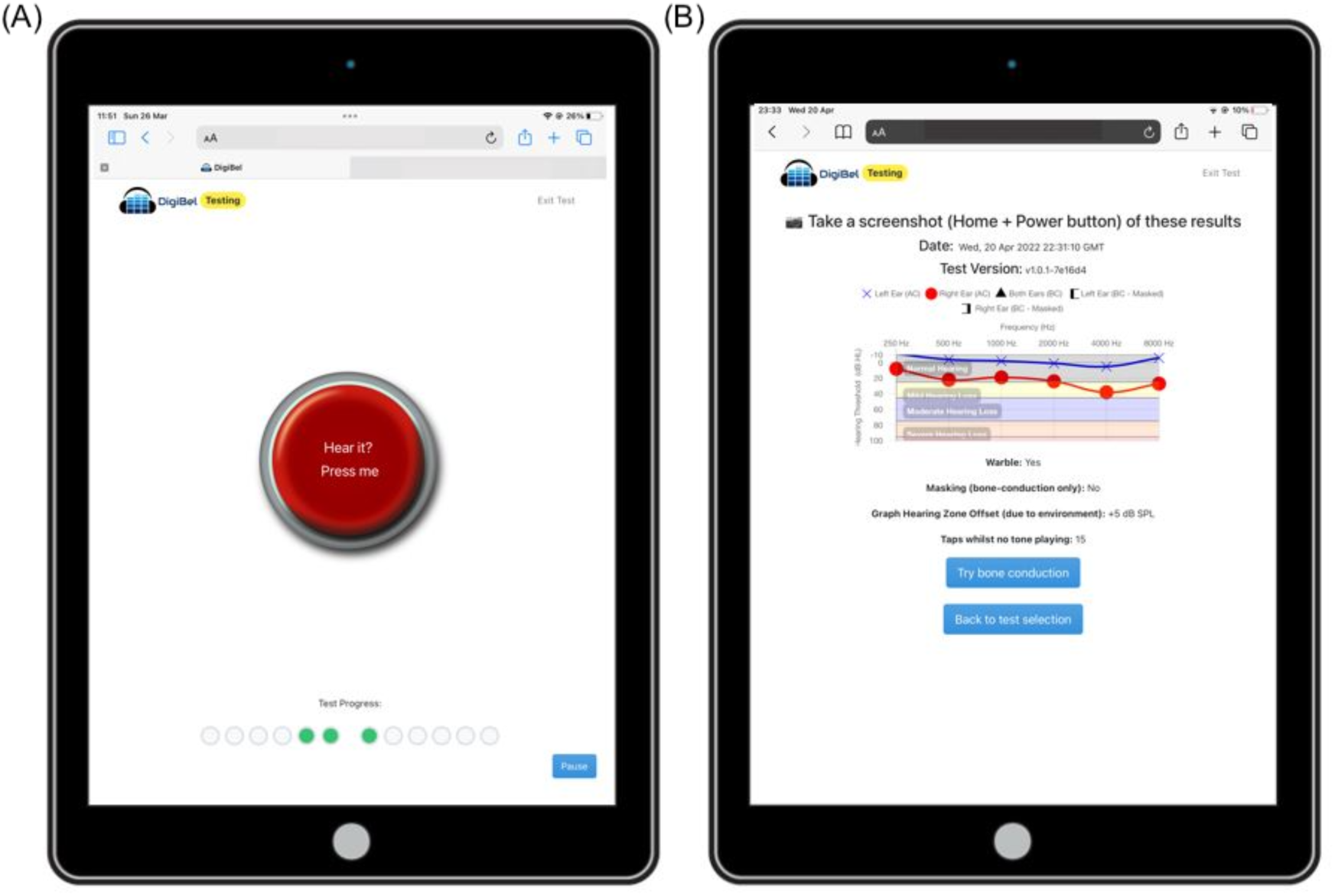
(A) DigiBel test interface (B) DigiBel audiometry graph

For this study, DigiBel AC PTA was undertaken in the sequence 2000, 4000, 8000, 1000, 500 and 250 Hz and retested in the sequence 2000, 4000, 8000, 500 Hz using the adult test version.

## Statistics

Data was analysed in R (version 4.1.2; R Foundation for Statistical Computing, Vienna, Austria). (11, 12) Accuracy and TRT reliability were assessed through Bland-Altman analysis and calculation of intraclass correlation coefficients (ICCs). Qualitative appraisal of ICC was based on conventional standards with moderate, good and excellent agreement indicated by ICC ≥0.50, ≥0.75 and ≥0.90 respectively. (13) The percentages of DigiBel threshold measurements lying within 10 dB of both the reference test and the repeated DigiBel test were calculated. (14) Statistical significance was calculated for the bias (mean difference between measurements) where confidence intervals did not cross zero, and for ICCs where *p* < 0.05. To assess diagnostic efficacy, sensitivity, and specificity for detection of hearing thresholds above 20Hz were calculated, using automated PTA as a reference. Student’s t-test was used to compare the number of false positive responses for each test. Throughout, magnitudes were reported as the mean (± standard deviation) unless otherwise stated.

## Results

### Participant characteristics

32 healthy subjects agreed to take part, but two were excluded due to malfunction of the reference automated PTA test. The 30 subjects who completed both the reference and index tests were aged 21-66 years of age (mean 27.9 ±10.3 years). TRT data was collected from 29 subjects (one volunteer left early due to time constraints). 24 subjects completed feedback forms (Fig 2).

**Fig 2.**
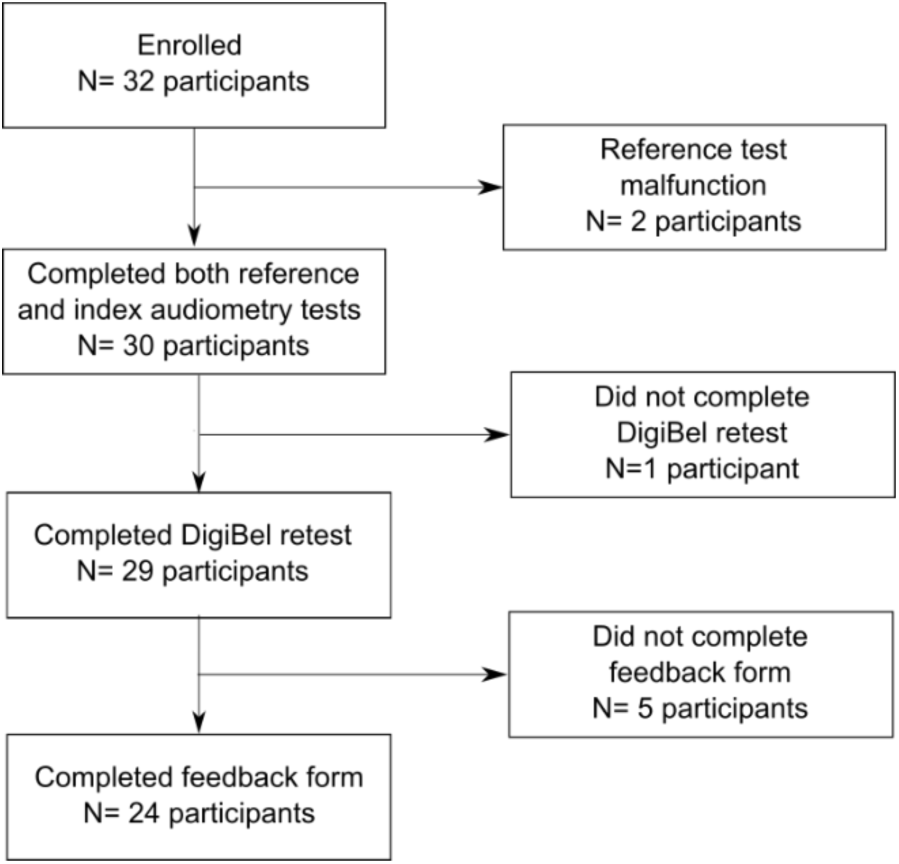
Participant flow diagram

### Accuracy and reliability of DigiBel

Across the six tested frequencies, threshold hearing levels of DigiBel compared to standard PTA gave ICC > 0.75 (good or excellent agreement) and *p* < 0.001 in every trial (Table 1).

**Table 1.** Comparison of threshold values in decibels (dB) using Bland Altman statistics and intra-class correlation coefficients (ICCs) between a) DigiBel and standard automated audiometry and b) DigiBel test and retest, where LLoA is the lower limit of agreement and ULoA is the upper limit of agreement.

| Frequency (Hertz) | N° Ears tested | Bias (dB)<br>(95% CI) | LLoA (dB)<br>(95% CI) | ULoA (dB)<br>(95% CI) | ICC<br>(95% CI) |
| --- | --- | --- | --- | --- | --- |
| **a) DigiBel compared to standard automated audiometry** |  |  |  |  |  |
| 250 | 60 | 1.72 ( -0.32 to 3.76) | -13.75 ( -17.25 to -10.25) | 17.18 (13.68 to 20.69) | 0.85 (0.77 to 0.91) |
| 500 | 60 | -1.55 ( -3.93 to 0.83) | -19.62 ( -23.71 to -15.52) | 16.52 (12.42 to 20.61) | 0.87 (0.80 to 0.92) |
| 1000 | 60 | 0.23 ( -2.30 to 2.77) | -19.01 ( -23.36 to -14.65) | 19.47 (15.11 to 23.83) | 0.88 (0.80 to 0.92) |
| 2000 | 60 | 2.43 ( -0.15 to 5.02) | -17.18 ( -21.62 to -12.74) | 22.05 (17.61 to 26.49) | 0.88 (0.81 to 0.93) |
| 4000 | 60 | 2.62 (0.14 to 5.10) | -16.19 ( -20.45 to -11.93) | 21.42 (17.16 to 25.68) | 0.89 (0.83 to 0.94) |
| 8000 | 60 | 4.60 (1.82 to 7.38) | -16.51 ( -21.29 to -11.73) | 25.71 (20.93 to 30.49) | 0.91 (0.82 to 0.95) |
| **b) DigiBel test-retest comparison** |  |  |  |  |  |
| 500 | 58 | 0.26 ( -1.91 to 2.43) | -15.90 ( -19.62 to -12.17) | 16.42 (12.69 to 20.14) | 0.92 (0.87 to 0.95) |
| 2000 | 58 | 1.40 ( -0.65 to 3.44) | -13.84 ( -17.36 to -10.33) | 16.64 (13.12 to 20.15) | 0.95 (0.91 to 0.97) |
| 4000 | 58 | 0.90 ( -0.66 to 2.46) | -10.74 ( -13.42 to -8.06) | 12.53 (9.85 to 15.22) | 0.97 (0.94 to 0.98) |
| 8000 | 58 | 0.98 ( -1.23 to 3.19) | -15.48 ( -19.28 to -11.68) | 17.44 (13.65 to 21.24) | 0.95 (0.92 to 0.97) |
| 8000 | 60 | 4.60 (1.82 to 7.38) | -16.51 ( -21.29 to -11.73) | 25.71 (20.93 to 30.49) | 0.91 (0.82 to 0.95) |
| **b) DigiBel test-retest comparison** |  |  |  |  |  |
| 500 | 58 | 0.26 ( -1.91 to 2.43) | -15.90 ( -19.62 to -12.17) | 16.42 (12.69 to 20.14) | 0.92 (0.87 to 0.95) |
| 2000 | 58 | 1.40 ( -0.65 to 3.44) | -13.84 ( -17.36 to -10.33) | 16.64 (13.12 to 20.15) | 0.95 (0.91 to 0.97) |
| 4000 | 58 | 0.90 ( -0.66 to 2.46) | -10.74 ( -13.42 to -8.06) | 12.53 (9.85 to 15.22) | 0.97 (0.94 to 0.98) |
| 8000 | 58 | 0.98 ( -1.23 to 3.19) | -15.48 ( -19.28 to -11.68) | 17.44 (13.65 to 21.24) | 0.95 (0.92 to 0.97) |

Mean lower limit of agreement (LOA) was -17.04 dB (±2.12 dB); mean upper LOA was 20.39 dB (±3.41 dB). No significant bias was apparent except at 4000 and 8000Hz where a small but statistically significant bias was apparent (2.62 and 4.60 dB, respectively), with DigiBel providing systematically higher threshold results at those frequencies (Fig 3). An agreeable level of threshold difference in audiometry assessments has previously been defined as 10dB; 73.1% of DigiBel threshold measurements were within this standard. (15)

**Fig 3.**
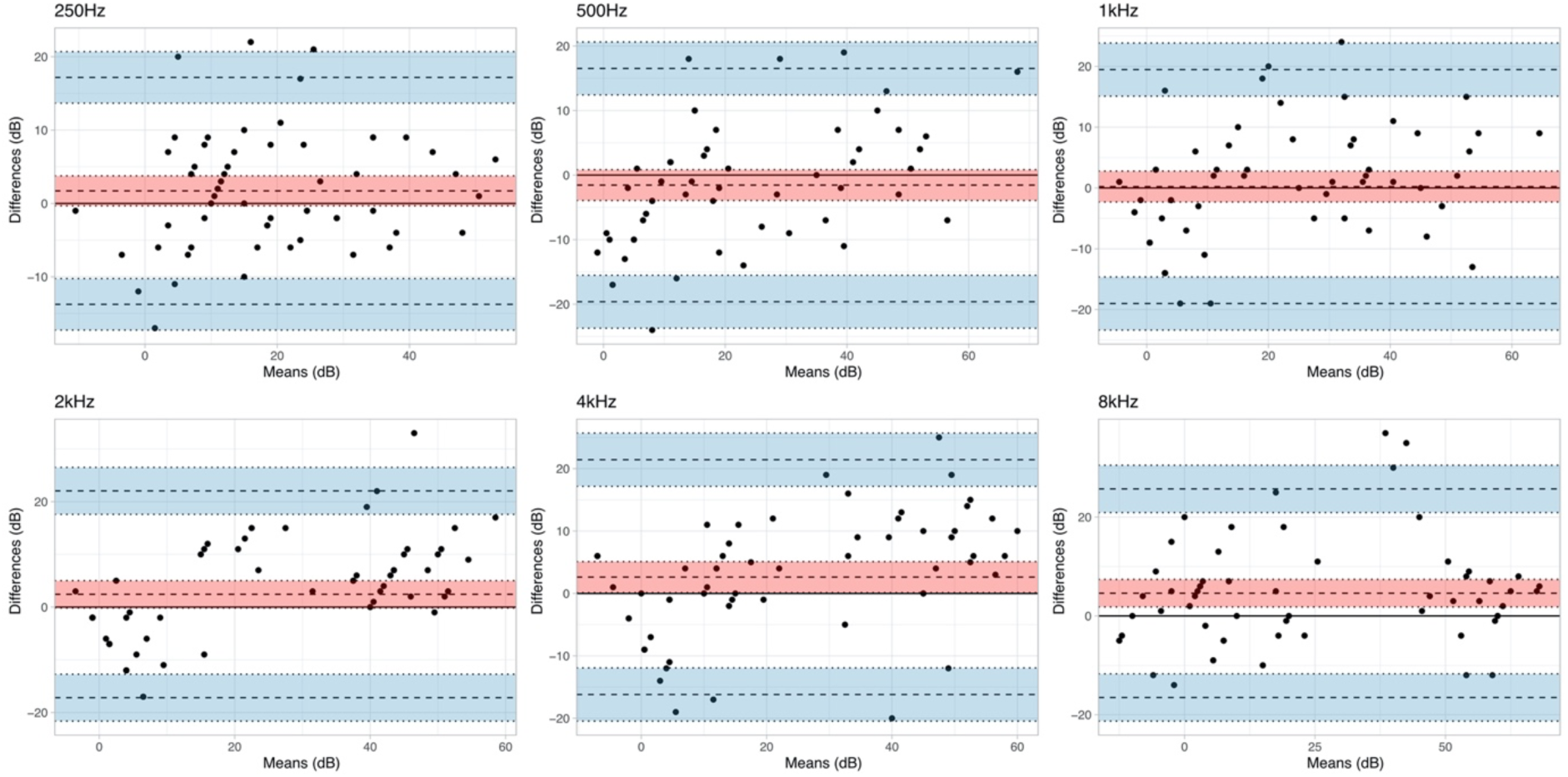
Bland-Altman plots comparing mean and difference in threshold measurements in decibels (dB) for DigiBel and standard automated pure tone audiometry at six frequencies (Hertz). 95% confidence intervals are shaded for the bias (red) and 95% limits of agreement (blue).

In TRT comparisons, ICC > 0.90 (excellent) at all tested frequencies: 500, 2000, 4000 and 8000Hz (Table 1). No statistically significant mean bias was exhibited at any frequency. Mean lower LOA was -13.99 dB (±2.34 dB); mean upper LOA was 15.76 dB (±2.20 dB). Overall, 84.6% of TRT thresholds were within 10dB of each other.

A mean 3.57±4.68 false positive responses were recorded during reference testing and 11.43±7.12 during the first DigiBel test (p<0.01).

### Sensitivity and specificity

The sensitivity and specificity of DigiBel for detecting 20dB hearing loss at each frequency is shown in Table 2. When applied to the four frequencies used for screening (250, 1000, 2000, 4000Hz), DigiBel had 100% sensitivity (95%CI 87.23-100) and 72.73% (95%CI 54.48-86.70) specificity for detecting 20dB hearing loss in adults in a quiet setting.

**Table 2.** Screening accuracy of DigiBel for hearing threshold >20 decibels (dB) identified by standard automated audiometry.

| Frequency (Hertz) | Sensitivity % (95%CI) | Specificity % (95%CI) |
| --- | --- | --- |
| 250 | 95.00 (75.13 to 99.87) | 85.00 (70.16 to 94.29) |
| 500 | 91.30 (71.96 to 98.93) | 89.19 (74.58 to 96.97) |
| 1000 | 100.00 (86.28 to 100.00) | 85.71 (69.74 to 95.19) |
| 2000 | 100.00 (86.77 to 100.00) | 79.41 (62.10 to 91.30) |
| 4000 | 100.00 (86.28 to 100.00) | 88.57 (73.26 to 96.80) |
| 8000 | 100.00 (86.28 to 100.00) | 88.57 (73.26 to 96.80) |
| Screening frequencies<br>(250,1000,2000, 4000) | 100.00 (87.23 to 100.00) | 72.73 (54.48 to 86.70) |

### Usability

21 of the 24 (87.5%) subjects completing the questionnaire did not regularly use digital health applications. All 24 rated DigiBel either good (62.5%) or excellent (37.5%); (7 (29%) preferred the DigiBel test, 6(25%) preferred the standard test and 11 (45.6%) gave no test preference. 21 (87.5%) agreed or strongly agreed that they would be confident to use DigiBel at home without help. The most common qualitative feedback given to the question “what is the best thing about the app?” was that it was easy and /or intuitive to use (7 (70.8%) of subjects). Answering “what is the worst thing about the app?” the commonest complaint was that the test was too long or boring (10 (41.7%) of subjects). One subject commented on environmental noise leak through the headphones.

## Discussion

At least five validated downloadable apps enable automated AC audiometry, several of which support self-testing without clinician involvement (16). Two, uHear and SHOEBOX for iOS, include a BC audiometry facility (17, 18). uHear has been calibrated for use with commercial in-ear Apple headphones and SHOEBOX uses purpose-built audiometry headphones. A potential advantage of DigiBel is that the system is calibrated to use affordable (retail price $41, €39), lightweight and wipe clean Sennheiser HD 400S AC headphones and Raspberry Pi BC headphones (retail price $28, €26).

DigiBel’s sensitivity and specificity for hearing impairment (>20dB threshold depression) screening was 100% (95%CI 87.2-100) and 72.7% (95%CI 54.45-86.7) respectively. This is comparable to previous studies of both uHear (98.2-100% sensitivity and 60.0-82.1 specificity) and SHOEBOX (91.2-93.3% and specificity of 57.8-94.5%). (18, 19, 17, 20, 21)

The hearing threshold measurement bias with DigiBel was insignificant compared to reference except at 4000 and 8000Hz, where the bias (2.62 and 4.60 dB higher than reference, respectively) reached statistical significance. Overall, 73% of threshold measurements with DigiBel were within 10dB of standard PTA; a previous study of SHOEBOX has found that 92.9% measurements were within this range. (14) The comparatively poor performance of DigiBel for this metric may be due to environmental noise leakage through the Sennheiser headphones, a disadvantage of their comfort.

Staircase methods of psychophysical threshold assessment are generally reported to be more reliable with less variability than ascending methods. (22) To our knowledge, DigiBel is the only application to apply this technique to audiometry, although it is commonly used for visual threshold testing. (13, 23) In this study, the number of false positive subject responses was significantly higher with DigiBel than standard testing in this study and may reflect its higher number of stimulus presentations compared to the ascending method used in standard automated PTA. The sensitivity of the iPad screen to a tap compared to the standard audiometer’s hand-held responder may an additional factor.

All subjects rated DigiBel good or excellent, but 41.7% complained that the test took too long or was boring. Test duration was not measured during this study, but threshold testing for six frequencies is expected to take approximately 13 minutes with DigiBel, several minutes longer than standard automated PTA, primarily due to its staircasing algorithm.

Study participants had also performed retests which may have contributed to perceived length of testing. Although the child-friendly version of the test is more entertaining, the test duration of threshold audiometry may limit its usability in young children. The DigiBel screening test of four frequencies takes approximately three minutes and may prove more feasible in this, its target population.

To simulate a range of hearing thresholds in the subject cohort, an earplug was used. This is a major limitation of the study because the effectiveness of the earplug may have altered during testing and earplugs may not accurately mimic genuine hearing impairment.

Although this study indicates that DigiBel is an acceptable and accurate method for detecting simulated hearing impairment in healthy adult volunteers, this is not generalisable to clinical settings.

This preliminary study confirms that the DigiBel app is an acceptable and easy-to-use self-testing online tool which accurately detects more than 20dB of simulated hearing impairment in adults. Future studies are planned to confirm DigiBel’s accuracy in clinical populations, particularly young children, and to determine if the app can identify those who may benefit from temporary use of a BC headphone / microphone kit at school, work and home while waiting for specialist care.

## Data Availability

All relevant data are within the manuscript.

## Acknowledgements

The authors thank Josephine Marriage and Martha Mann at Chears-audiology, Royston, Cambridge for their support with biologic calibration of the DigiBel app and Tamsin Holland-Brown for improving the manuscript.

## Conflicts of interest

Louise Allen is the inventor of the DigiBel app and founding director of Cambridge Medical Innovation Ltd.

## Funding

This study received no funding.

## Notes

### Competing Interest Statement

Louise Allen is the inventor of the DigiBel app and founding director of Cambridge Medical Innovation Ltd. We confirm that these competing interests will not alter adherence to PLOS policies on sharing data and materials.

### Author Declarations

1. Non-abbreviated, full names and affiliations of all ethics oversight bodies that ruled on ethics of your study - Patient Outcomes Department, Safety and Quality Support of Cambridge University Hospitals NHS Foundation Trust 2. Decision made, i.e. whether ethical approval was given or waived - “This project has been reviewed and approved with the Patient Outcomes Department and confirmed as a Service Evaluation. This project has been approved via a 4 stage approval process by the projects Audit Lead, Specialty Audit Lead, Directorate Audit Lead and Clinical Audit Coordinator for this Specialty/Directorate. This project was reviewed and approved with no ethical review required.” Signed by Mrs Amy Baker (Patient Outcomes Manager, Cambridge University Hospitals NHS Foundation Trust)

## References

1. World report on hearing. Geneva: World Health Organization; 2021.

2. Williamson IG, Dunleavey J, Bain J, Robinson D. The natural history of otitis media with effusion--a three-year study of the incidence and prevalence of abnormal tympanograms in four South West Hampshire infant and first schools. J Laryngol Otol 1994; 108(11):930–4.

3. Lusignan S de, Correa A, Pathirannehelage S, Byford R, Yonova I, Elliot AJ et al. RCGP Research and Surveillance Centre Annual Report 2014-2015: disparities in presentations to primary care. Br J Gen Pract 2017; 67(654):e29–e40.

4. Leach AJ, Homøe P, Chidziva C, Gunasekera H, Kong K, Bhutta MF et al. Panel 6: Otitis media and associated hearing loss among disadvantaged populations and low to middleincome countries. Int J Pediatr Otorhinolaryngol 2020; 130 Suppl 1(Suppl 1):109857.

5. UK National Screening Committee. Child Screening Programme: Hearing (Child). Available from: URL: https://view-health-screening-recommendations.service.gov.uk/hearing-child/.

6. Prieve BA, Schooling T, Venediktov R, Franceschini N. An Evidence-Based Systematic Review on the Diagnostic Accuracy of Hearing Screening Instruments for Preschool- and School-Age Children. Am J Audiol 2015; 24(2):250–67.

7. Fonseca S, Forsyth H, Neary W. School hearing screening programme in the UK: practice and performance. Arch Dis Child 2005; 90(2):154–6.

8. Holland Brown TM, Fitzgerald O’Connor I, Bewick J, Morley C. Bone conduction hearing kit for children with glue ear. BMJ Innov 2021; 7(4):600–3.

9. Carhart R JJ. Preferred methods for clinical determination of pure-tone thresholds. J.Speech Hear. Res. 1959; 24:330–45.

10. British Society of Audiology. Recommended Pure Tone Audiometry guidelines. Available from: URL: https://www.thebsa.org.uk/wp-content/uploads/2018/11/OD104-32-Recommended-Procedure-Pure-Tone-Audiometry-August-2018-FINAL-1.pdf.

11. Wickham H, Averick M, Bryan J, Chang W, McGowan L, François R et al. Welcome to the Tidyverse. JOSS 2019; 4(43):1686.

12. Tsagris M, Papadakis M. Forward Regression in R: From The Extreme Slow to the Extreme Fast. Journal of Data Science 2018; 16(4):771–80.

13. Thirunavukarasu AJ, Mullinger D, Rufus-Toye RM, Farrell S, Allen LE. Clinical validation of a novel web-application for remote assessment of distance visual acuity. Eye (Lond) 2022; 36(10):2057–61.

14. Bastianelli M, Mark AE, McAfee A, Schramm D, Lefrançois R, Bromwich M. Adult validation of a self-administered tablet audiometer. J Otolaryngol Head Neck Surg 2019; 48(1):59.

15. Lemkens N, Vermeire K, Brokx JPL, Fransen E, van Camp G, van de Heyning PH. Interpretation of pure-tone thresholds in sensorineural hearing loss (SNHL): a review of measurement variability and age-specific references. Acta Otorhinolaryngol Belg 2002; 56(4):341–52.

16. Bright T, Pallawela D. Validated Smartphone-Based Apps for Ear and Hearing Assessments: A Review. JMIR Rehabil Assist Technol 2016; 3(2):e13.

17. Yeung JC, Heley S, Beauregard Y, Champagne S, Bromwich MA. Self-administered hearing loss screening using an interactive, tablet play audiometer with ear bud headphones. Int J Pediatr Otorhinolaryngol 2015; 79(8):1248–52.

18. Szudek J, Ostevik A, Dziegielewski P, Robinson-Anagor J, Gomaa N, Hodgetts B et al. Can Uhear me now? Validation of an iPod-based hearing loss screening test. J Otolaryngol Head Neck Surg 2012; 41 Suppl 1:S78–84.

19. Mahomed F, Swanepoel DW, Eikelboom RH, Soer M. Validity of automated threshold audiometry: a systematic review and meta-analysis. Ear Hear 2013; 34(6):745–52.

20. Abu-Ghanem S, Handzel O, Ness L, Ben-Artzi-Blima M, Fait-Ghelbendorf K, Himmelfarb M. Smartphone-based audiometric test for screening hearing loss in the elderly. Eur Arch Otorhinolaryngol 2016; 273(2):333–9.

21. Handzel O, Ben-Ari O, Damian D, Priel MM, Cohen J, Himmelfarb M. Smartphone-based hearing test as an aid in the initial evaluation of unilateral sudden sensorineural hearing loss. Audiol Neurootol 2013; 18(4):201–7.

22. Running CA. High false positive rates in common sensory threshold tests. Atten Percept Psychophys 2015; 77(2):692–700.

23. Johnson CA, Chauhan BC, Shapiro LR. Properties of staircase procedures for estimating thresholds in automated perimetry. Invest Ophthalmol Vis Sci 1992; 33(10):2966–74.

